# Tryptophan and arginine metabolism is significantly altered at the time of admission in hospital for severe COVID-19 patients: findings from longitudinal targeted metabolomics analysis

**DOI:** 10.1101/2021.03.31.21254699

**Authors:** Laura Ansone, Monta Ustinova, Anna Terentjeva, Ingus Perkons, Liga Birzniece, Vita Rovite, Baiba Rozentale, Ludmila Viksna, Oksana Kolesova, Kristaps Klavins, Janis Klovins

## Abstract

The heterogeneity in severity and outcome of COVID-19 cases points out the urgent need for early molecular characterization of patients followed by risk-stratified care. The main objective of this study was to evaluate the fluctuations of serum metabolomic profiles of COVID-19 patients with severe illness during the different disease stages in a longitudinal manner. We demonstrate a distinct metabolomic signature in serum samples of 32 hospitalized patients at the acute phase compared to the recovery period, suggesting the tryptophan (tryptophan, kynurenine, and 3-hydroxy-DL-kynurenine) and arginine (citrulline and ornithine) metabolism as contributing pathways in the immune response to SARS-CoV-2 with a potential link to the clinical severity of the disease. In addition, we provide evidence for glutamine metabolism in M2 macrophages as a complementary process and contribution of phenylalanine and tyrosine in the molecular mechanisms underlying the severe course of the infection. In conclusion, our results provide several functional metabolic markers for disease progression and severe outcome with potential clinical application.

**Importance:** Although the host defense mechanisms against SARS-CoV-2 infection are still poorly described, they are of central importance in shaping the course of the disease and the possible outcome. Metabolomic profiling may complement the lacking knowledge of the molecular mechanisms underlying clinical manifestations and pathogenesis of COVID-19. Moreover, early identification of metabolomics□based biomarker signatures is proved to serve as an effective approach for the prediction of disease outcome. Here we provide the list of metabolites describing the severe, acute phase of the infection and bring the evidence of crucial metabolic pathways linked to aggressive immune responses. Finally, we suggest metabolomic phenotyping as a promising method for developing personalized care strategies in COVID-19 patients.

## Main text

More than a year has passed since the World Health Organization (WHO) announced the COVID-19 outbreak as a pandemic in March 2020, following the rapid spread of the SARS-CoV-2 virus (1). The clinical course of COVID-19 is versatile, the infection of the SARS-CoV-2 virus not only varies in its severity from asymptomatic or mild and moderate respiratory disease (80%) to clinically severe or critical life-threatening disease (20%) but also varies in a range of organs the disease can affect (2, 3). Diverse clinical trajectories seem to be the result of the immune response differences between individuals (4).

Multiple innate and adaptive immune system pathways that produce inflammatory molecules against the virus and virus-infected human cells are triggered after the SARS-CoV-2 entry in the cell, with characteristic overexpression of proinflammatory cytokines (e.g. IL-6, TNFα, IFN-γ) known as cytokine storm in the most severe cases (4–6). The host’s immune responses typically involve changes in metabolic processes at the cellular level, reflecting the host-defense mediators and underlying mechanisms (7).

Detailed understanding of the molecular mechanisms behind COVID-19 pathogenesis and inflammatory response is needed to predict and reduce individual risks, develop therapeutic strategies, and reduce the overall ∼2% mortality rate (mortality in hospitalized patients can be up to 30%) (8, 9). The human blood sera metabolome (defined as small molecules <1500–2000 Da) reflects the organism’s metabolic state and is widely used to gain a deeper understanding of the pathogenesis of diseases. Recent reports of metabolomics studies highlight the pivotal role of cellular metabolites in programming immune response to SARS-CoV-2 infection (10–13). Considering the extremely high heterogeneity of the COVID-19 disease and lack of promising predictive biomarkers, we believe that implications of longitudinal metabolite profiling may be beneficial in understanding the underlying mechanisms of the diverse course of the disease and promote the early identification of people at increased risk of severe illness from COVID-19 and related complications.

We performed quantitative targeted metabolome analysis with liquid chromatography-mass spectrometry (LC-MS) in blood sera of 32 hospitalized COVID-19 patients at the acute phase (time of admission at the hospital) and the recovery phase (40 ± 14.92 days) of the disease (see Text S1 in the supplemental material for a detailed description of methods). Written informed consent was obtained from every participant before their inclusion in the study, and the study protocol was approved by the Central Medical Ethics Committee of Latvia (No. 01-29.1.2/928).

As expected, the clinical blood tests revealed abnormal hematological parameters for the majority of study participants at the time of hospitalization, with a high variation in platelet (202.94 ± 65.26 µL) levels and low lymphocyte measurements (0.64 ± 0.56 µL), which coincides with previously reported lymphopenia as the hallmark of severe COVID-19 cases. We also observed a high variation of several markers (e.g. alanine aminotransferase, bilirubin, lactate C-reactive protein) indicating renal and hepatic dysfunction, myocarditis, inflammation, and coagulation, which confirms the systemic response to the infection in our study cohort(2, 14) (see Table S1 in the supplemental material).

Out of 51 metabolites analyzed by LC-MS, 22 metabolites showed significantly altered levels (paired t-test, FDR<0.05) in the serum samples during the acute phase in comparison to the recovery phase (Table 1), where concentrations for 16 compounds were significantly elevated, whereas 8 metabolites were decreased. The hierarchical clustering and principal component analysis of the obtained metabolomic profiles showed clear metabolomics-based discrimination of samples collected in different phases of the disease (Figure1. A and B), indicating an altered metabolic activity during infection. Pathway analysis revealed 13 significantly enriched pathways (FDR<0.05), including phenylalanine, tyrosine and tryptophan biosynthesis, D-glutamine and D-glutamate metabolism and arginine biosynthesis (Figure1 C, Table S3). Statistical analysis was done with Metaboanalyst version 5.0 (15).

**Table 1.**
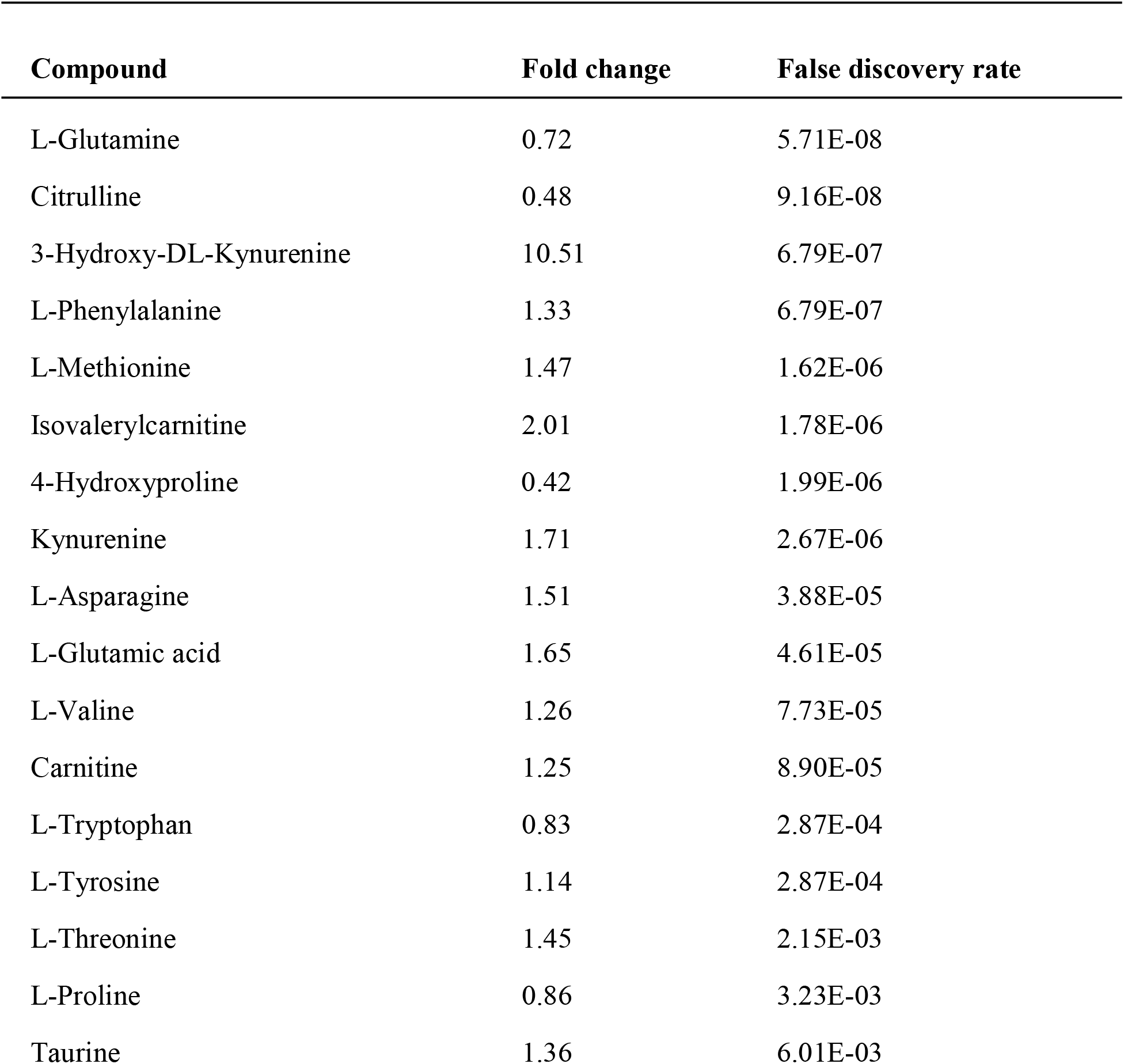
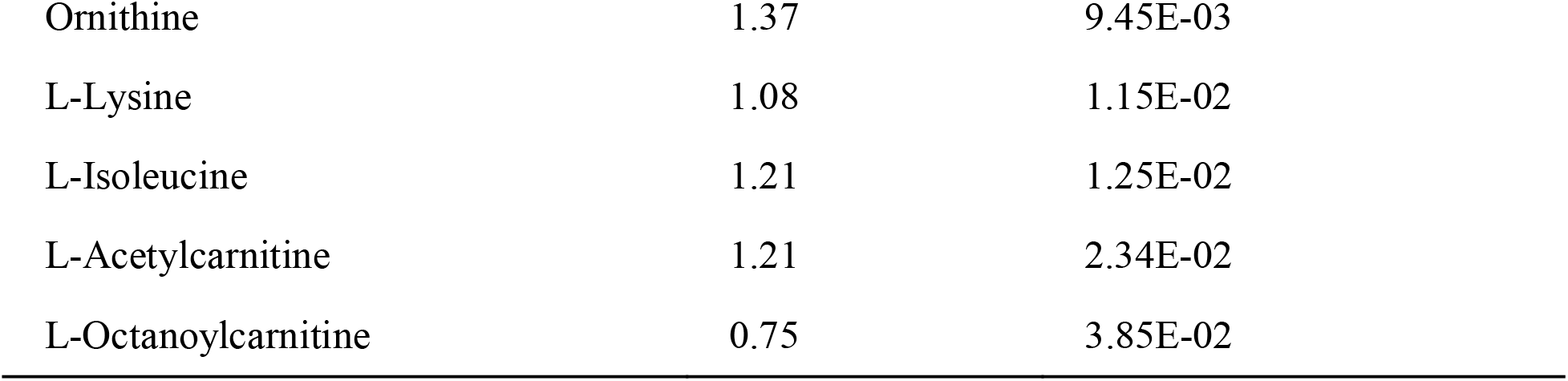
Serum metabolites showing significantly altered levels comparing measures obtained during the acute phase and recovery phase of the disease.

**Figure 1.**
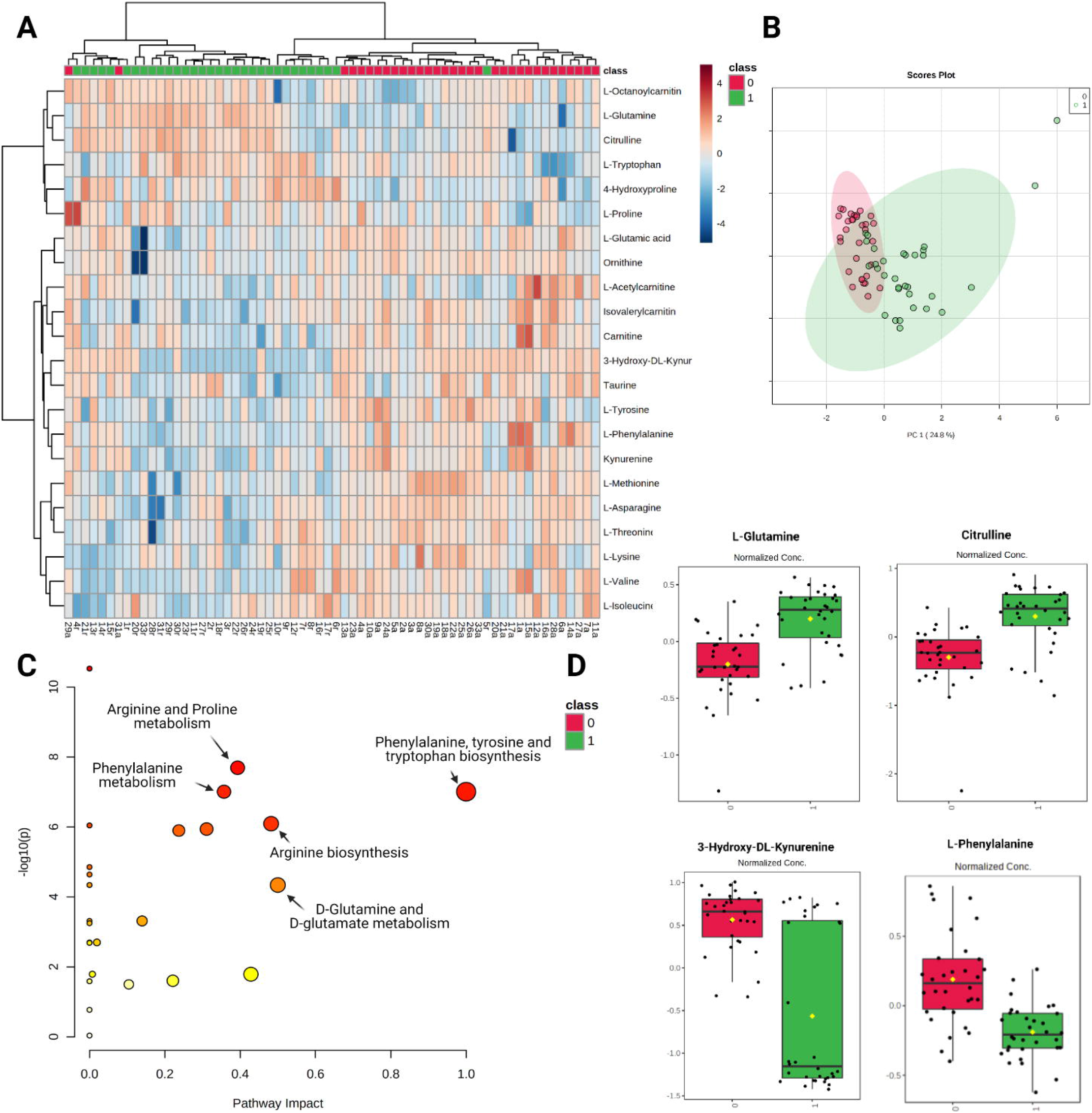
Targeted metabolomic analysis of longitudinal serum samples of hospitalized COVID-19 patients. (A) Heatmap and hierarchical clustering of top 22 significantly altered metabolites. Each column represents one sample: (0, green) - samples collected in the acute phase, (1, red) - samples collected during the recovery phase, each row conforms to a specific metabolite expressed in normalized, log transformed concentration value. (B) Principal component analysis showing clear discrimination of samples collected in the acute and recovery phases of infection based on the obtained metabolite profiles. (C) Scatter plot representing the most relevant metabolic pathways from KEGG library arranged by adjusted p-values (obtained by Global Test pathway enrichment analysis) on Y-axis, and pathway impact values (from pathway topology analysis) on X-axis. The node color is based on its p-value and the node radius is determined based on their pathway impact values. (D) Boxplots showing the normalized levels of the most functionally relevant metabolites during the acute phase (0) and the recovery phase (1) of the infection, described as the minimum value, the first quartile, the median, the third quartile, and the maximum value with the black dots representing each sample.

We found L-glutamine (Figure 1D) as the most significantly changed amino acid between the paired samples with reduced acute phase concentrations. It is known that glutamine deprivation and decreased glutaminolysis inhibits M2 macrophage polarization, which may partly explain the hyperinflammatory state in severe COVID-19 cases (16, 17). Moreover, the beneficial effect of glutamine has been proposed in multiple studies, where adding enteral L-glutamine to the regular nutrition shortened the duration of hospitalization and improved the outcome in moderate to severe COVID-19 cases (18, 19).

Two other amino acids involved in arginine catabolism through the Urea cycle: citrulline (Figure 1D) and ornithine (Table 1) were found to significantly change between the acute and recovery phases. However, alterations in the levels of L-arginine itself were not detected. Low blood plasma citrulline levels have already been reported in COVID-19 patients, whereas in patients with severe sepsis, the decreased citrulline levels are associated with acute respiratory distress syndrome (11, 13, 20). Since arginine can be metabolized to creatine and then to creatinine, both varying highly in our study cohort according to regular blood tests performed during hospitalization, arginine catabolism may be implicated in disturbed kidney function of COVID-19 patients (21).

Three out of 22 metabolites showing significantly changed levels between acute infection and recovery phase are involved in the tryptophan-kynurenine pathway: L-tryptophan, kynurenine, and 3-hydroxy-DL-kynurenine (Figure 1D, Table 1). The reduction of tryptophan levels and increase of kynurenine, 3-hydroxy-DL-kynurenine in the acute phase supports the conclusions of previous reports and confirms this pathway’s key role in severe COVID-19 cases (10, 11). *In vitro* experiments have shown that tryptophan deprivation sensitizes T cells to apoptosis, inhibits proliferation of T cells, and plays a role in CD8 T-cell suppression in cancer (22, 23). Notably, the tryptophan-kynurenine pathway shows a modulatory effect on the macrophage-mediated responses by targeting the synthesis of the metabolic coenzyme NAD+(24, 25). According to Thomas *et al*. dysregulated tryptophan metabolism, an essential regulator of inflammation and immunity may be a potential explanation for severity in older COVID-19 patients (Thomas et al., 2020).

Finally, we observed a significant difference in L-phenylalanine (Figure 1D) and tyrosine levels in our cohort between the analyzed disease phases, both already suggested as metabolic hot spots of COVID-19 before (26). In sepsis and HIV-1 infection, the increased phenylalanine sera concentrations are linked to immune activation and increased cardiovascular event risk (27– 29). Although the mechanisms behind this association are not well studied, it is in line with microvascular endothelial damage and higher coagulation risk characteristic to both: coronary heart disease and severe COVID-19 (29). Phenylalanine and tyrosine are catabolized to dopamine and epinephrine, and the latter has been employed in cardiac arrest as a result of cytokine storm, characteristic to severe COVID-19 patients(30).

In conclusion, our study shows that metabolomic profiling provides novel insights into the pathogenesis of host-defense mechanisms and may be further applied for rapid biomarker discovery in infectious disease. These discoveries could show novel therapeutic strategies as indirect targets to fasten the recovery process after severe COVID-19. To the best of our knowledge, this is the first longitudinal study covering metabolomic profiling of severe COVID-19 patients.

## Data Availability

The raw data supporting the current study are available from the corresponding author on request.

## Data availability

The raw data supporting the current study are available from the corresponding author on request.

## Acknowledgments

This study is funded by the Ministry of Education and Science, Republic of Latvia, project “Establishment of COVID-19 related biobank and integrated platform for research data in Latvia”, project No. VPP-COVID-2020/1-0016.

We acknowledge The Boris and Inara Teterev Foundation for support to Riga Stradins University in patient sample collection.

The authors acknowledge the Latvian Biomedical Research and Study Centre and the Genome Database of the Latvian Population for providing the infrastructure, biological material, and data.

## Author contributions

LA, KK, IP, LB performed the sample analysis; MU, VR, LA, BR, LV, OK were involved in patient recruitment process and sample collection; AT ensured the clinical data acquisition; MU, LA drafted the manuscript; JK provided critical revision of the manuscript; LA, KK, MU performed the data analysis and interpretation; MU, JK collaborated in funding acquisition, JK performed supervision and conceptualization of the study. All authors read and approved the final manuscript.

## Appendixes

Text S1. Detailed Methods Description.

Table S1. Characteristics of the study participants.

Table S2. Data matrix with quantified metabolite concentrations for targeted compounds, determined by ultra-performance liquid chromatography-mass spectrometry.

Table S3. Significant pathways based on enrichment procedures.

## References

1. World Health Organization. 2020. Coronavirus disease 2019 (COVID-19) Situation Report – 51.

2. Zhou F, Yu T, Du R, Fan G, Liu Y, Liu Z, Xiang J, Wang Y, Song B, Gu X, Guan L, Wei Y, Li H, Wu X, Xu J, Tu S, Zhang Y, Chen H, Cao B. 2020. Clinical course and risk factors for mortality of adult inpatients with COVID-19 in Wuhan, China: a retrospective cohort study. Lancet 395:1054–1062.

3. Wu Z, McGoogan JM. 2020. Characteristics of and Important Lessons from the Coronavirus Disease 2019 (COVID-19) Outbreak in China: Summary of a Report of 72314 Cases from the Chinese Center for Disease Control and Prevention. JAMA - J Am Med Assoc 323:1239–1242.

4. García LF. 2020. Immune Response, Inflammation, and the Clinical Spectrum of 11:4–8.

5. Qin C, Zhou L, Hu Z, Zhang S, Yang S, Tao Y, Xie C, Ma K, Shang K, Wang W, Tian DS. 2020. Dysregulation of immune response in patients with coronavirus 2019 (COVID- 19) in Wuhan, China. Clin Infect Dis 71:762–768.

6. Mehta P, McAuley DF, Brown M, Sanchez E, Tattersall RS, Manson JJ. 2020. COVID-19: consider cytokine storm syndromes and immunosuppression. Lancet 395:1033–1034.

7. Pearce EL, Pearce EJ. 2013. Metabolic Pathways in Immune Cell Activation and Quiescence. Immunity 38:633–643.

8. Asch DA, Sheils NE, Islam N, Chen Y, Werner RM, Buresh J, Doshi JA. 2020. Variation in US Hospital Mortality Rates for Patients Admitted With COVID-19 During the First 6 Months of the Pandemic 19104:1–8.

9. Bellan M, Patti G, Hayden E, Azzolina D, Pirisi M, Acquaviva A, Aimaretti G, Valletti PA, Angilletta R, Arioli R, Avanzi GC, Avino G, Balbo PE, Baldon G, Baorda F, Barbero E, Baricich A, Barini M, Adesi FB, Battistini S, Beltrame M, Bertoli M, Bertolin S, Bertolotti M, Betti M, Bobbio F, Boffano P, Boglione L, Borrè S, Brucoli M, Calzaducca E, Cammarata E, Cantaluppi V, Cantello R, Capponi A, Carriero A, Casciaro FG, Castello LM, Ceruti F, Chichino G, Chirico E, Cisari C, Cittone MG, Colombo C, Comi C, Croce E, Daffara T, Danna P, Corte F Della, Vecchi S De, Dianzani U, Benedetto D Di, Esposto E, Faggiano F, Falaschi Z, Ferrante D, Ferrero A, Gagliardi I, Gaidano G, Galbiati A, Gallo S, Garavelli PL, Gardino CA, Garzaro M, Gastaldello ML, Gavelli F, Gennari A, Giacomini GM, Giacone I, Via VG, Giolitti F, Gironi LC, Gramaglia C, Grisafi L, Inserra I, Invernizzi M, Krengli M, Labella E, Landi IC, Landi R, Leone I, Lio V, Lorenzini L, Marzari L, Marzullo P, Mennuni M, Montabone C. 2020. Fatality rate and predictors of mortality in an Italian cohort of hospitalized COVID LJ 19 patients. Sci Rep 1–10.

10. Blasco H, Bessy C, Plantier L, Lefevre A, Piver E, Bernard L, Marlet J, Stefic K, Benz-de Bretagne I, Cannet P, Lumbu H, Morel T, Boulard P, Andres CR, Vourc’h P, Hérault O, Guillon A, Emond P. 2020. The specific metabolome profiling of patients infected by SARS-COV-2 supports the key role of tryptophan-nicotinamide pathway and cytosine metabolism. Sci Rep 10:1–12.

11. Thomas T, Stefanoni D, Reisz JA, Nemkov T, Bertolone L, Francis RO, Hudson KE, Zimring JC, Hansen KC, Hod EA, Spitalnik SL, D’Alessandro A. 2020. COVID-19 infection alters kynurenine and fatty acid metabolism, correlating with IL-6 levels and renal status. JCI Insight 5.

12. Shen B, Yi X, Sun Y, Bi X, Du J, Zhang C, Quan S, Zhang F, Sun R, Qian L, Ge W, Liu W, Liang S, Chen H, Zhang Y, Li J, Xu J, He Z, Chen B, Wang J, Yan H, Zheng Y, Wang D, Zhu J, Kong Z, Kang Z, Liang X, Ding X, Ruan G, Xiang N, Cai X, Gao H, Li L, Li S, Xiao Q, Lu T, Zhu YJ, Liu H, Chen H, Guo T. 2020. Proteomic and Metabolomic Characterization of COVID-19 Patient Sera. SSRN Electron J https://doi.org/10.1101/2020.04.07.20054585.

13. Lawler NG, Gray N, Kimhofer T, Boughton B, Gay M, Yang R, Morillon A-C, Chin S-T, Ryan M, Begum S, Bong SH, Coudert JD, Edgar D, Raby E, Pettersson S, Richards T, Holmes E, Whiley L, Nicholson JK. 2021. Systemic Perturbations in Amine and Kynurenine Metabolism Associated with Acute SARS-CoV-2 Infection and Inflammatory Cytokine Responses. J Proteome Res https://doi.org/10.1021/acs.jproteome.1c00052.

14. Henry BM, Helena M, Oliveira S De, Benoit S. 2020. Hematologic, biochemical and immune biomarker abnormalities associated with severe illness and mortality in coronavirus disease 2019 (COVID-19): a meta-analysis 58:1021–1028.

15. Xia J, Wishart DS. 2011. Web-based inference of biological patterns, functions and pathways from metabolomic data using MetaboAnalyst. Nat Protoc 6:743–760.

16. Liu P-S, Wang H, Li X, Chao T, Teav T, Christen S, Di Conza G, Cheng W-C, Chou C-H, Vavakova M, Muret C, Debackere K, Mazzone M, Huang H-D, Fendt S-M, Ivanisevic J, Ho P-C. 2017. α-ketoglutarate orchestrates macrophage activation through metabolic and epigenetic reprogramming. Nat Immunol 18:985–994.

17. Martinez FO, Combes TW, Orsenigo F, Gordon S. 2020. Monocyte activation in systemic Covid-19 infection: Assay and rationale. EBioMedicine 59:102964.

18. Cengiz M, Uysal BB, Ikitimur H, Ozcan E, Aktepe E, Yavuzer H, Yavuzer S. 2020. Effect of oral L -Glutamine supplementation on Covid-19 treatment 33:24–31.

19. Obayan AOE. 2021. Infectious Diseases and Epidemiology Overview of the Rationale for L-Glutamine Treatment in Moderate-Severe COVID-19 Infection Proposed Flowchart of In fl ammatory Response In COVID-19 Infec D on 7:1–7.

20. Ware LB, Magarik JA, Wickersham N, Cunningham G, Rice TW, Christman BW, Wheeler AP, Bernard GR, Summar ML. 2013. Low plasma citrulline levels are associated with acute respiratory distress syndrome in patients with severe sepsis 1–8.

21. Reyes AA, Karl IE, Klahr S. 1994. Role of arginine in health and in renal disease. Am J Physiol Physiol 267:F331–F346.

22. Lee GK, Park HJ, Macleod M, Chandler P, Munn DH, Mellor AL. 2002. Tryptophan deprivation sensitizes activated T cells to apoptosis prior to cell division. Immunology 107:452–460.

23. Greene LI, Bruno TC, Christenson JL, D’Alessandro A, Culp-Hill R, Torkko K, Borges VF, Slansky JE, Richer JK. 2019. A Role for Tryptophan-2,3-dioxygenase in CD8 T-cell Suppression and Evidence of Tryptophan Catabolism in Breast Cancer Patient Plasma. Mol Cancer Res 17:131 LP – 139.

24. Minhas PS, Liu L, Moon PK, Joshi AU, Dove C, Mhatre S, Contrepois K, Wang Q, Lee BA, Coronado M, Bernstein D, Snyder MP, Migaud M, Majeti R, Mochly-Rosen D, Rabinowitz JD, Andreasson KI. 2019. Macrophage de novo NAD+ synthesis specifies immune function in aging and inflammation. Nat Immunol 20:50–63.

25. Van Gool F, Gallí M, Gueydan C, Kruys V, Prevot P-P, Bedalov A, Mostoslavsky R, Alt FW, De Smedt T, Leo O. 2009. Intracellular NAD levels regulate tumor necrosis factor protein synthesis in a sirtuin-dependent manner. Nat Med 15:206–210.

26. Pang Z, Zhou G, Chong J, Xia J. 2021. Comprehensive meta-analysis of covid-19 global metabolomics datasets. Metabolites 11:1–14.

27. Zangerle R, Kurz K, Neurauter G, Kitchen M, Sarcletti M, Fuchs D. 2010. Increased blood phenylalanine to tyrosine ratio in HIV-1 infection and correction following effective antiretroviral therapy. Brain Behav Immun 24:403–408.

28. Ploder M, Neurauter G, Spittler A, Schroecksnadel K, Roth E, Fuchs D. 2008. Serum phenylalanine in patients post trauma and with sepsis correlate to neopterin concentrations. Amino Acids 35:303–307.

29. Würtz P, Havulinna AS, Soininen P, Tynkkynen T, Prieto-Merino D, Tillin T, Ghorbani A, Artati A, Wang Q, Tiainen M, Kangas AJ, Kettunen J, Kaikkonen J, Mikkilä V, Jula A, Kähönen M, Lehtimäki T, Lawlor DA, Gaunt TR, Hughes AD, Sattar N, Illig T, Adamski J, Wang TJ, Perola M, Ripatti S, Vasan RS, Raitakari OT, Gerszten RE, Casas J-P, Chaturvedi N, Ala-Korpela M, Salomaa V. 2015. Metabolite profiling and cardiovascular event risk: a prospective study of 3 population-based cohorts. Circulation2015/01/08. 131:774–785.

30. Luo P, Liu D, Li J. 2021. Epinephrine use in COVID-19: friend or foe? Eur J Hosp Pharm 28:e1 LP–e1.

