## Supplementary table 1 for "Tryptophan and arginine metabolism is significantly altered at the time of admission in hospital for severe COVID-19 patients: findings from longitudinal targeted metabolomics analysis"

Table S1. Characteristics of the study participants.

| **Variable** | **Mean (SD) or n (%)** |
|---|---|
| Gender (male/female) | 16 (48.5 %)/17 (51.5 %) |
| Age (years) | 58.52 (16.26) |
| BMI (kg/m$^2$) | 32.66 (24.04) |
| Time spent in hospital (days) | 11.67 (5.22) |
| **Comorbidities** | |
| **Number of patients with comorbidities, (yes/no)** | 21 (63.6 %) /12 (36.4%) |
| Hypertension | 15 (45.5%) |
| Type 2 diabetes mellitus | 3 (9.1%) |
| Respiratory system disease | 1 (3.3%) |
| Cardiovascular disease | 7 (21.2%) |
| Malignancies | 1 (3.3%) |
| Other disease | 10 (30.3 %) |
| **Biochemical measurements** | |
| Leukocytes (µL) | 5.52 (1.66) |
| Hemoglobin (g/dL) | 13.74 (1.27) |
| Hematocrit (%) | 41.41 (3.56) |
| Platelets (µL) | 202.94 (65.26) |
| Neutrophils (µL) | 2.18 (1.78) |
| Lymphocytes (µL) | 0.64 (0.56) |
| Monocytes (µL) | 0.24 (0.20) |
| Eosinophils (µL) | 0.02 (0.04) |
| ALT (U/l) | 33.78 (25.41) |
| ASAT (U/l) | 41.94 (27.24) |
| GGT (U/l) | 79.09 (86.25) |
| Bilirubin (µmol/l) | 9.19 (5.54) |
| LDH (U/L) | 312.46 (177.19) |
| Potassium (mmol/L) | 4.10 (0.42) |
| Sodium (mmol/L) | 134.67 (4.13) |
| Creatinine (µmol/L) | 80.47 (28.66) |
| CRP (mg/L) | 56.98 (58.43) |
| D-dimer (mg/ml) | 83.89 (256.23) |
| Ferritin (mg/L) | 720.55 (1149.92) |

| | |
|---|---|
| Troponin T (pg/mL) | 13.11 (10.33) |

SD: standard deviation; BMI: body mass index; ALT: alanina aminotranferase; AST: aspartate aminotransferase; GGT: gamma-glutamyl transferase; LDH: lactate dehydrogenase; CRO: C-reactive protein.
