## Supplementary table 3 for "Tryptophan and arginine metabolism is significantly altered at the time of admission in hospital for severe COVID-19 patients: findings from longitudinal targeted metabolomics analysis"

Table S3. Significant pathways based on enrichment procedures.

| **Pathway Name** | **False discovery rate** | **Impact** |
|---|---|---|
| Phenylalanine, tyrosine and tryptophan biosynthesis | 7.14E-07 | 1 |
| D-Glutamine and D-glutamate metabolism | 9.50E-05 | 0.5 |
| Arginine biosynthesis | 3.76E-06 | 0.48223 |
| Taurine and hypotaurine metabolism | 0.020547 | 0.42857 |
| Arginine and proline metabolism | 2.95E-07 | 0.39328 |
| Phenylalanine metabolism | 7.14E-07 | 0.35714 |
| Alanine, aspartate and glutamate metabolism | 4.09E-06 | 0.3109 |
| Tryptophan metabolism | 4.09E-06 | 0.23722 |
| Histidine metabolism | 0.028892 | 0.22131 |
| Tyrosine metabolism | 8.84E-04 | 0.13972 |
| Cysteine and methionine metabolism | 0.034102 | 0.10446 |
| Glutathione metabolism | 0.0028641 | 0.01966 |
| Primary bile acid biosynthesis | 0.020547 | 0.00758 |
