## Supplementary text 1 for "Tryptophan and arginine metabolism is significantly altered at the time of admission in hospital for severe COVID-19 patients: findings from longitudinal targeted metabolomics analysis"

### Detailed Methods Description

#### Study Design and patient recruitment

In this study, we performed the longitudinal serum metabolome analysis in 33 Covid-19 patients with severe course of the disease. Patient recruitment was conducted in the major hospitals of Latvia (Riga East University Hospital and Vidzeme Hospital) during the acute phase of the disease. Written informed consent was obtained from every participant before their inclusion in the study, and the study protocol was approved by the Central Medical Ethics Committee of Latvia (No. 01-29.1.2/928). The peripheral blood samples for this study were collected in 1-3 days after admission to the hospital (acute phase) and 40.30±14.92 days later (recovery phase). Biochemical blood tests were performed in 0-48 hours from the hospitalization moment, clinical data were collected from the medical records of each hospital. A longitudinal study design was chosen as the most suitable approach for evaluating metabolomics data characterized with high inter-individual variability.

#### Sample processing

Serum separation was performed by centrifuging peripheral blood samples collected in BD Vacutainer Blood Collection tubes at 4000 rpm, +4 C for 15 minutes. 800 µL of methanol was added to 200 µL of serum samples. Samples were vortexed for 10 seconds, shaken for 20 min at 450 rpm, and centrifuged for 10 min at 10000 g. 100 μL of sample extract was dried down using a centrifugal vacuum evaporator. Dry residues were reconstituted in 200 μL of methanol and 20 μL of the isotopically labeled internal standard mix was added. Afterward, samples were transferred to HPLC vials and used for LC-MS analysis.

**LC-MS analysis**

A Dionex 3000 HPLC system (Thermo Scientific) coupled with an Orbitrap Q Exactive (Thermo Scientific) mass spectrometer was used for the LC-MS analysis. The chromatographic separation was carried out on an ACQUITY UPLC BEH Amide, 1.7 μm, 2.1x100 mm analytical column (Waters) equipped with a VanGuard: BEH C18, 2.1x5 mm pre-column (Waters). The column was maintained at a temperature of 40 °C and the sample injection volume was 2 μL. The mobile phase consisted of phase A - 0.15% formic acid (v/v) in water and phase B - 0.15% formic acid (v/v) in 85% acetonitrile (v/v) with 10 mM ammonium formate. The gradient elution with a flow rate of 0.4 mL/min was performed for a total analysis time of 17 min. The Orbitrap Q Exactive (Thermo Scientific) mass spectrometer was operated in a positive electrospray ionization mode, spray voltage 3.5 kV, aux gas heater temperature 400 °C, capillary temperature 350 °C, aux gas flow rate 12, sheat gas flow rate 50. The metabolites of interest were analyzed using a full MS scan mode, scan range m/z 50 to 400, resolution 35000, AGC target 1e6, maximum IT 50 ms. The Trace Finder 4.1 software (Thermo Scientific) was used for data processing. A seven-point linear calibration curve with internal standardization and 1/x weighing was constructed for the quantification of the metabolites.

### Data analysis

Data matrix containing time point information and quantified metabolite concentrations (µmol/L) that were above Level Of Detection (LOD) in more than 50% of the samples were uploaded in web-based, free software Metaboanalyst, version 5.0 (https://www.metaboanalyst.ca/). The uploaded data file contained 66 samples (33 pairs) by 31 compounds data matrix. A total of 67 (3.3%) missing values were detected, missing values were replaced by 1/5 of the min positive values of their corresponding variables. One of the samples was identified as an outlier and both samples from that patient were excluded from further analysis. Normalization by sum, log transformation, and Pareto scaling was performed. The univariate analysis included paired Fold Change analysis to evaluate the levels and direction of change of metabolites and paired t-test to identify significant targeted metabolites between acute and recovery phase. Principal Component Analysis (PCA) was performed to evaluate the distribution of samples within 95% confidence region, the two outliers in PCA were due to significantly low L-Cystine level in two patients in recovery phase sera samples. The heatmap was generated with normalized data and 22 significant features from the t-test, using Ward Clustering Algorithm and Euclidean Distance Measure. The pathway analysis was also done in Metaboanalyst 5.0 (https://www.metaboanalyst.ca/), quantified concentrations (µmol/L) were uploaded, data were normalized by sum and log transformation, Pareto scaling was applied. Pathway analysis was visualized with Scatter plot, Enrichment method was set to Global Test and as Reference pathway library was selected *Homo Sapiens* (KEGG). The figures were drawn via metaboanalyst software v 5.0 (https://www.metaboanalyst.ca/). For average, percentage, and standard deviation calculations in phenotype data *MS Excel* was used.
